# Artificial intelligence to predict the risk of mortality from Covid-19: Insights from a Canadian Application

**DOI:** 10.1101/2020.09.29.20201632

**Authors:** Brett Snider, Paige Phillips, Aryn MacLean, Edward McBean, S. Andrew Gadsden, John Yawney

## Abstract

The Severe Acute Respiratory Syndrome COVID-19 virus (SARS-CoV-2) has had enormous impacts, indicating need for non-pharmaceutical interventions (NPIs) using Artificial Intelligence (AI) modeling. Investigation of AI models and statistical models provides important insights within the province of Ontario as a case study application using patients’ physiological conditions, symptoms, and demographic information from datasets from Public Health Ontario (PHO) and the Public Health Agency of Canada (PHAC). The findings using XGBoost provide an accuracy of 0.9056 for PHO, and 0.935 for the PHAC datasets. Age is demonstrated to be the most important variable with the next two variables being Hospitalization and Occupation. Further, AI models demonstrate identify the importance of improved medical practice which evolved over the six months in treating COVID-19 virus during the pandemic, and that age is absolutely now the key factor, with much lower importance of other variables that were important to mortality near the beginning of the pandemic.

An XGBoost model is shown to be fairly accurate when the training dataset surpasses 1000 cases, indicating that AI has definite potential to be a useful tool in the fight against COVID-19 even when caseload numbers needed for effective utilization of AI model are not large.

## I. Introduction

THE Severe Acute Respiratory Syndrome COVID-19 virus (SARS-CoV-2) has had enormous impacts throughout the world, spreading at an alarming rate such that there have now been more than 28.6 million infections and 917 thousand deaths globally, including >6 million cases and 193 thousand deaths in the United States alone (Johns Hopkins, 2020). The world is collectively frustrated by the limited ability to predict and control future changes in the virus trajectory. The virus is pervasive, resulting in the need to develop a better understanding how alternative non-pharmaceutical interventions (NPIs) can be implemented to control the worldwide outbreak (including, but not limited to, shutdown of economies, and lockdown of peoples’ movements), while keeping the virus under a controllable magnitude until a vaccine becomes available.

Mammoth changes in the operations of economies and populations have arisen as most countries have adopted self-isolation as a means of decreasing infectivity levels and holding down the curve (‘planking’ or ‘flattening’ the curve of the virus). While successful in some countries, other nations continue to evolve into resurgence of the virus as a result of ‘opening up the economy’ too quickly (e.g. the USA) or unable to enact social distancing and extensive poverty (e.g. India). These examples emphasize the need for insights on how governments need a guide for allowable behavior controls and means of opening up the economy as society moves forward.

To provide insights into the effectiveness of alternative initiatives, the power of Artificial Intelligence (AI) as a technology is investigated as a means to improve prediction of the future impact on the medical care system. Internationally, and more specifically within Canada, the pandemic has caused widespread lockdowns on activities within economies. Further, enormous angst arose in New York city, as an example, where medical preparedness was pushed to the limit during the height of their pandemic, where ventilators to assist people with breathing problems proved to be in short supply and were under frequent re-distribution between cities within the US. These types of implications questioning whether available Personal Protective Equipment (PPE) resources were expressed, as the virus raged through various country populations. President Trump undertook efforts to restrict forwarding of PPE outside the US, much to the concern of foreign countries including Canada, where agreements had been established, and hence, represented an attempt to derail forwarding along of PPE materials; these types of actions will change the attitudes in terms of reliance on foreign countries to provide critical items at times of need (the capacities of medical systems are definitely limited).

Concerns continue, including issues of a potential resurgence of virus infections if removal of lockdown conditions occurs too early and a second wave of the virus commences. If too early an opening of the economy occurs, or relaxation of some measures such as opening daycare or summer day camps for children, these actions may increase the spread of the virus. Some states, such as Florida, Texas and Arizona, opened up their economies very early and were then confronted with the difficult task of trying to reimplement lockdown procedures in hopes of controlling the rapidly spreading virus.

There continues to be concern that Canada’s hospital system will become overwhelmed, and this possibility underlines the need for forecasting models to provide the country’s leaders with information (e.g., how many people are likely going to be admitted to hospitals, and how many will require intensive care and/or ventilators, who is most at risk, etc.) to help them make informed decisions. In response to the highly complex nature of the COVID-19 outbreak and variations in its behavior internationally, machine learning has potential to model the viral outbreak [1].

In response, and to improve the understanding of the success of various initiatives related to COVID-19 attenuation, this paper describes research related to the development of AI and statistical models that provide innovative insights into the COVID-19 pandemic. This research was undertaken to compare alternative models regarding their ability to identify the mortality risk of COVID-19 patients in the province of Ontario. Data-based methodologies were implemented to enable widespread usage of the resultant model, adaptable to various public health units in their planning for inflated caseloads on medical systems as well as to inform the public about who is at greatest risk. This research was undertaken to create city-wide and region-wide models aimed at improving predictions of mortality within the province of Ontario.

Although the applications of AI modelling of the COVID-19 outbreak are still in their infancy, this paper demonstrates significant benefits of evolving these models to improve mortality predictions from COVID-19, and hospital caseloads within a province of Canada context. This paper is organized as follows. The use of machine learning to address COVID-19 is discussed in Section 2. The case study and problem setup are discussed in Section 3. Section 4 includes the experimental results and discussion of applying machine learning to study COVID-19. The paper is then concluded.

## II. Machine Learning to Address COVID-19

Machine Learning, as an approach to AI, enables improved predictions of the future impact on the healthcare system, stemming from the overall effect of the virus on the general population. In response to the highly complex nature of the COVID-19 outbreak and variations in its behavior internationally, researchers suggest machine learning is an effective tool to model the viral outbreak [1]. More precisely, multiple machine learning methods can be used to analyse COVID-19 cases in Ontario, with case information prevailing from two different publicly available datasets. These methods include Artificial Neural Network (ANN), Extreme Gradient Boosting (XGBoost), and Random Forest. Logistic Regression, a statistical regression-based model was also used for the predictions in this paper so that the Machine Learning results can be compared to the results of a simpler model. Other applications of the aforementioned Machine Learning algorithms have proven to be valuable in an enormous array of applications, including just as examples, landslide susceptibility models, predicted price changes in crude oil, and geographical origin analysis of music samples, respectively [2, 3, 4].

The ability to make reasonably accurate predictions on the future outcomes of COVID-19 have the potential to have an immense impact on choices surrounding behavioural directives, as well as decisions made for Canada’s healthcare system. Even with some degrees of self-isolation and social distancing, conditions are indicating a real possibility that Canada’s hospital system may become overwhelmed [5]. To provide an acceptable response to the demand for hospital beds, it is imperative that an approach is developed to distinguish between high risk and low risk COVID-19 patients.

### A. AI Modelling in Healthcare

The role of AI in healthcare is being increasingly well established over the past decade. The insights available from interpreting diverse and vast datasets can produce lifesaving results. The advanced integration and instant access of technology allows for better management of patient data, spanning from the security and administration of electronic medical records, effortless linkage of data sources, and real-time information sharing. This elevated level of data collection consequently leads to improved data manipulation. However, to effectively make use of the large and complex datasets including in-depth health data, there is need for epidemiologists and health care professionals to incorporate computational techniques that identify patterns in data [6]. This reinforces the need for use of Machine Learning and other mathematical modelling as key analytical tools in healthcare epidemiology [7].

Studies have shown that AI can be a valuable tool in the modelling and prediction of viral infections within the population. A 2018 study effectively mapped the transmission of the mosquito-born Zika virus (ZIKV) using multiple Machine Learning models; Backward Propagation Neural Network (BPNN), Gradient Boosting Machine (GBM) and Random Forest [8]. These models were used to generate the probability of Zika outbreak at the global level, with an area under the curve (AUC) between 0.963 and 0.966, indicating a very high level of accuracy [8]. This application of Machine Learning for the 2015 Zika epidemic established that AI methods are effective for disease transmission prediction and risk assessment.

Machine Learning tools are not only useful for modelling the spread of a virus, but can also predict patient outcomes. In a study supported by the U.S. National Institute on Aging, researchers generated a ‘super learning model’, an ensemble of multiple Machine Learning approaches, that collectively forms a predictive algorithm with the best cross-validated mean-squared-error [9]. The findings concluded that Machine Learning methods for epidemiological predictions are highly dependent on typical characteristics such as gender and age, plus demonstrating accuracy can be augmented with additional biostatistics such as smoking habits, underlying heart conditions, reported physical activity tendencies, level of education, household income, and weight [9].

The various Machine Learning algorithms used in the study [11], were also integrated within a 2019 forecast that monitored vital signs of Intensive Care Unit patients, and whether or not they would proceed to critical conditions [10]. Further, accuracy was found in the Random Forest Machine Learning algorithm for an analysis for predicting patient conditions remotely, improving existing single predictive models [11].

### B. Early COVID-19 Results

Specific to COVID-19 results, public health authorities, as well as common media, are placing attention on traditional epidemiological and statistical models. Although quick to develop with fewer datasets and foci [12], it has already been determined that standard models fitted to confirmed cases of COVID-19 are anticipated to have a high level of uncertainty, largely due to the deviation on reported dates for confirmed cases [13]. Moreover, the popularized Susceptible-Exposed-Infectious-Recovered (SEIR) model, as well as the simpler, SIR model, can both provide useful insights into the effectiveness of transmission prevention strategies, but can still be expected to derive results with considerable uncertainty [14].

AI modelling of the pandemic has had good preliminary results. A study focussed on the development of AI predictions as a form of decision-making support had up to 80% accuracy when using a K-Nearest Neighbors algorithm [15]. Surprisingly, this study found that characteristics of COVID-19, including fever, certain features in radiographic lung images, and strong immune responses were not useful in predicting which of the sampled 53 patients (all evidencing with initially mild symptoms) would progress to develop severe lung disease. Age and gender were also ineffective factors in predicting the development of serious disease. Instead, the results indicated that a combination of small changes in levels of the liver enzyme alanine aminotransferase (ALT), patient-reported myalgia, and raised hemoglobin levels most accurately predicted the development of serious disease conditions.

Researchers at China’s Huazhong University of Science and Technology have used Machine Learning (using XGBoost) to develop a prognostic prediction algorithm to forecast the likelihood of an individual surviving the infection [16].

Overall, there are a number of primary areas of scope where AI has demonstrated significant potential in fighting COVID-19, including early warnings and alerts, tracking and prediction, data dashboards, diagnosis and prognosis, treatments, and cures, and societal control. However, the utility of AI models depends on the availability of pertinent data. Specifically, although AI models will not replace the role of experts or trusted epidemiological models, AI models have the potential to monitor and respond to the crisis – assisting in the quantification of medical caseloads and future global trends [17]. In a whitepaper by *ttopstart*, it was outlined that, regardless of the challenges in acquiring the necessary data and integrating the algorithms within existing medical workflows, AI will likely make immense differences in the future of healthcare administration and delivery [18].

### C. Adapting AI Models for Ontario, Canada

AI models for Ontario as described herein, were developed to assist hospitals and healthcare facilities to decide which patients to prioritize in receiving medical attention, elevate to hospitalization, or triage when the system is overwhelmed. The algorithm predicts the mortality risks based on patients’ physiological conditions, symptoms, and demographic information. These data were collected from two sources, Public Health Ontario (PHO) and the Public Health Agency of Canada (PHAC), both of which cluster datasets by geographic health regions. In Ontario, these 36 regionalized health authorities are called Public Health Units, as depicted in Figure 1. Due to large clusters of cases, Ontario is of great interest for this application of AI modeling.

**Figure 1.**
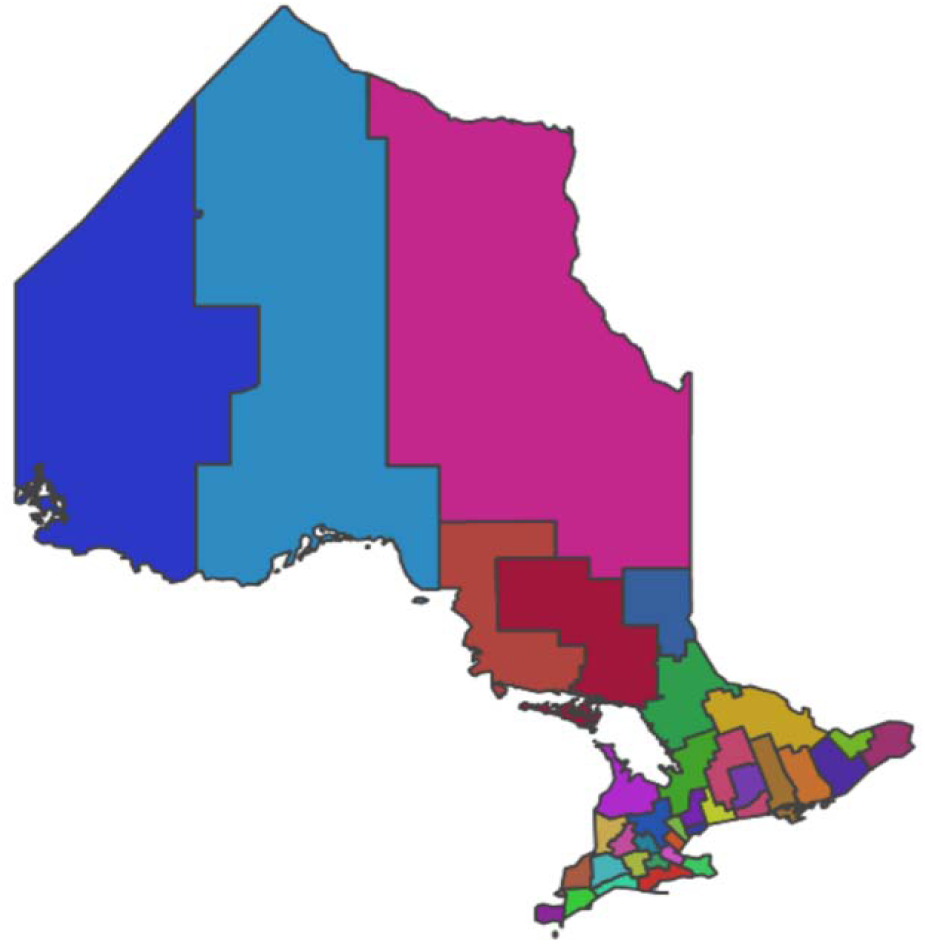
Public Health Units groups by geographical health regions in the Province of Ontario [19].

## III. Case Study and Computer Experiments

Cumulatively, the available data across all regions of Ontario was for 39,794 individual cases (August 1, 2020), either grouped under recovery or mortality. One dimension of these outbreaks, considered the largest, was within the Province’s long-term care facilities. Between January 15 and June 1, 2020, nearly 18% of all provincial COVID-19 cases were confirmed as long-term care residents [20]. During this time, rates of COVID-19 within long-term care per 100,000 population reached as high as 72.8, as reported wit in the Leeds, Grenville & Lanark District Health Unit [20].

This research developed and contrasted various machine learning and statistical models in order to predict the risk of mortality for Canadians infected with COVID-19. The models were calibrated using only publicly accessible Canadian COVID-19 data to highlight the applicability of AI modeling, Specifically, two open access COVID-19 dataset with varying attribute information, were used to build and compare the models. The first dataset (PHO) was obtained from Ontario Health Services [21], while the second (PHAC) was obtained from Canada health services [22]. To ensure fair comparisons were made between datasets, the COVID-19 cases were limited to only Ontario COVID-19 cases that were reported after January 19, 2020 and were resolved (recovered or died) by Aug 1, 2020. A summary of the two datasets and their attributes are listed below in Table 1.

**TABLE I.** Characteristic of Ontario and Canadian Individual COVID-19 Case Data.

| Variable | Ontario Dataset (PHO) | Canadian Dataset (PHAC) |
| --- | --- | --- |
| Age | Categorical: (<20, 20-29, 30-39... 90+) | Categorical |
| Public Health Unit | Categorical (all 33 public health units in Ontario) | Not Available |
| Date Infected | Numeric (Days from Jan 1,2020 to expected date of infection) | Numeric (Weeks from Jan 1,2020 to expected date of infection or earliest known date of infection) |
| Date Reported | Numeric (Days from Jan 1,2020 to reported date) | Not Available |
| Gender | Categorical (M, F, Other) | Categorical (M, F, Other) |
| Case Acquisition | Categorical (close contact, community spread, outbreak, travel, Unknown) | Categorical (Domestic acquisition, International travel, Unknown) |
| Occupation | Not Available | Categorical (healthcare worker, other) |
| Hospitalization | Not Available | Categorical (ICU, hospital, neither) |
| Cases Recovered | 36,988 | 27,778 |
| Cases Died | 2806 | 2612 |

The Machine Learning models were adapted for Ontario to focus on three distinct case statuses, namely active, recovered, or reported deceased. PHO data reports 36, 988 cases between January 19^th^ and August 1^st^ [23], PHAC report 27,778 cases in Ontario for the same period [24]. Data after August 1^st^ was not used as the caseloads prior to this were already substantial. Detailed characteristics of the website information in terms of variable definitions for both the PHO and PHAC datasets are as described in Table 1.

As evident in Table 1, several variables are the same across both datasets (Gender), similar but with minor differences (age, date infected, case acquisition), and several variables are only present in one or the other (Public Health Unit, Date Reported, Occupation and Hospitalization). By comparing the accuracy of models built using the different publicly available datasets, the importance of specific variables can be highlighted as well as begin to predict how a more complete dataset may improve the models’ accuracy.

Also worth noting, is that the number of COVID-19 cases that recovered or died between Jan 23-Aug 16, 2020 differ between datasets by 194 deaths. This highlights that data anomalies and/or differences in reporting requirements exist between the two datasets. More detailed description of the reporting requirements and methodology for each dataset would be useful to identify what is causing the difference in COVID-19 cases but with the enormous speed of the pandemic, data assembly challenges continue to exist.

### A. Model Selection

Three machine learning models and one statistical regression model were developed to predict the mortality risk of COVID-19 patients within Ontario. The models selected include; artificial neural network (ANN) [25], Random Forest [26], extreme gradient boosting decision tree (XGBoost) [27], and logistic regression [25]. These models were selected based on their prevalence in literature and accuracy in binary classification.

### B. Data Processing

Before calibrating any of the models, the datasets were randomly divided into two parts; training dataset containing 70% of the data, and the testing dataset containing the remaining 30% of the data. The training dataset was used to calibrate each model, whereas the testing dataset was used to evaluate the final models’ accuracy.

Pre-processing techniques were applied to the training dataset. First, numeric variables were centered and scaled, and categorical variables were converted into dummy (or binary) variables. The SMOTE resampling technique was applied to the training dataset to adjust for the class imbalance (i.e. only a small number of COVID-19 patients actually die from this disease) [28].

Tuning parameters for the machine learning models (ANN, Random Forest, XGBoost) were determined using a grid search approach and assessed using a 10-fold cross-validation technique repeated three times for each model. Tuning parameters were optimized to produce the maximum area under the receiver operating characteristic curve (Area Under the Curve, or AUC) based on the average 10-fold cross-validation technique.

The logistic regression’s input variables were selected using a step-wise Akaike Information Criterion (AIC) function [25], which selects the array of input variables to be used based on step-wise selection process which is evaluated using the AIC.

All models were developed and analyzed using the computer programming language R [29]. The models generate a probability of mortality for each recorded case outcome, and then decide the final predicted outcome, be it recovered or died, based on a 50% threshold value.

## IV. Experimental Results And Discussion

The AUC is an effective indicator measurement of accuracy to represent the performance of a classification model. In general, the AUC represents the degree to which the model can distinguish between different classes [30], and in this case, the classes being either a patient has recovered or died. It is the area under the Receiver Operating Characteristics (ROC) curve, with 1.0 being a perfect measure of separability and 0.0 means that the model is only generating false predictions [31]. When the AUC is near 0.5, it means that the model cannot recognise the different possible classes and predicts outcomes at random.

The AUCs of each model for both datasets are depicted below in Figure 2.

**Figure 2.**
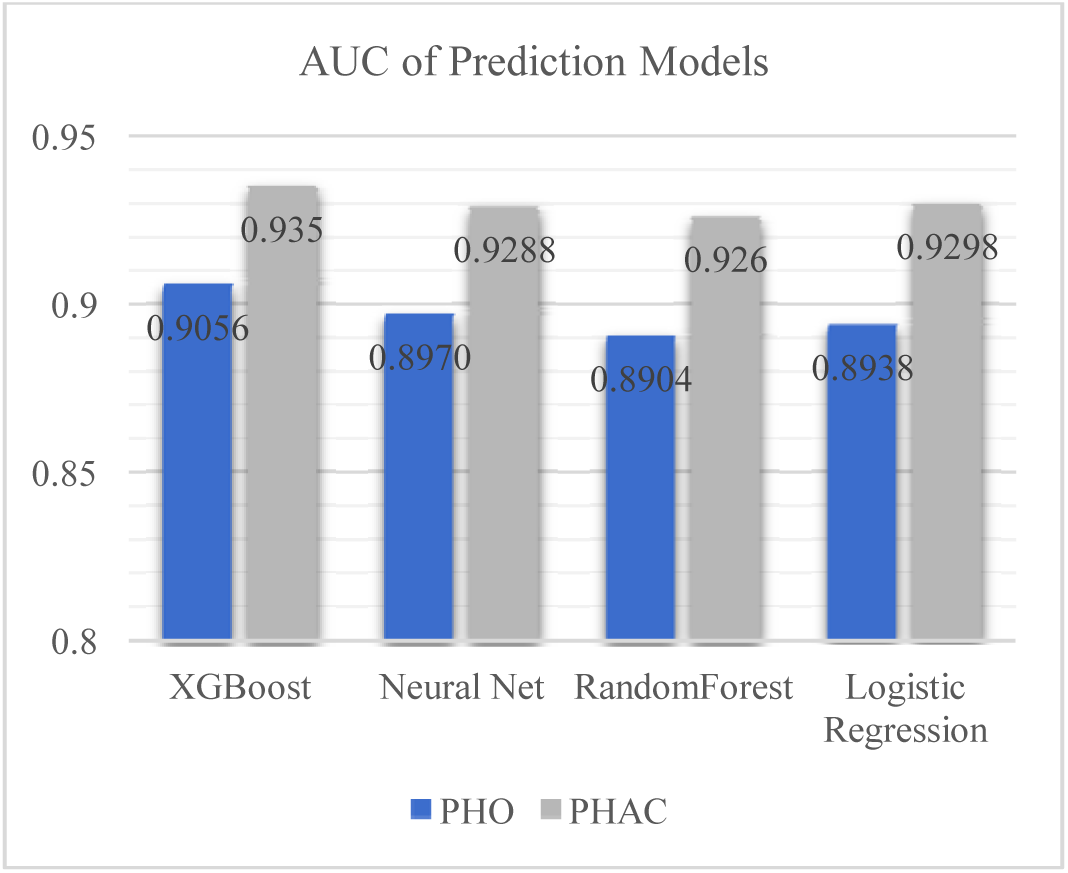
Area Under the Curve of mortality prediction models for both the PHO and PHAC datasets.

Figure 2 highlights that for both datasets, all models have relatively high AUC values (>0.88). This indicates all models are able to distinguish which patients are most likely to recover and which ones have the highest risk of death with a high degree of accuracy. The most accurate model was XGBoost, achieving an AUC of 0.9056 for PHO and 0.935 for the PHAC datasets.

The Logistic Regression model has the second-highest AUC for both PHO and PHAC datasets. This is interesting since regression models are often found to be less accurate than machine learning models, especially when datasets contain a large number of input variables, although in this application, since there are only six variables, the advantage of machine learning models ability to identify complex relationships between input variables and the outcomes are somewhat limited. These findings indicate that not being able to incorporate the complex relationships between input variables and the outcome is important but not terribly deficient, when compared to machine learning models, as the logistic model produces results better than ANN and Random Forest.

### A. Variable importance between datasets

As illustrated in Figure 2, all models developed using the PHAC dataset outperformed the models developed using the PHO dataset. This indicates the variables included in the PHAC dataset provide greater information in predicting COVID-19 mortality, than the variables included in the PHO dataset. To characterize this further, a variable importance graph is provided in Figure 3, indicating importance of individual variables for the most accurate model in both datasets (XGBoost).

**Figure 3.**
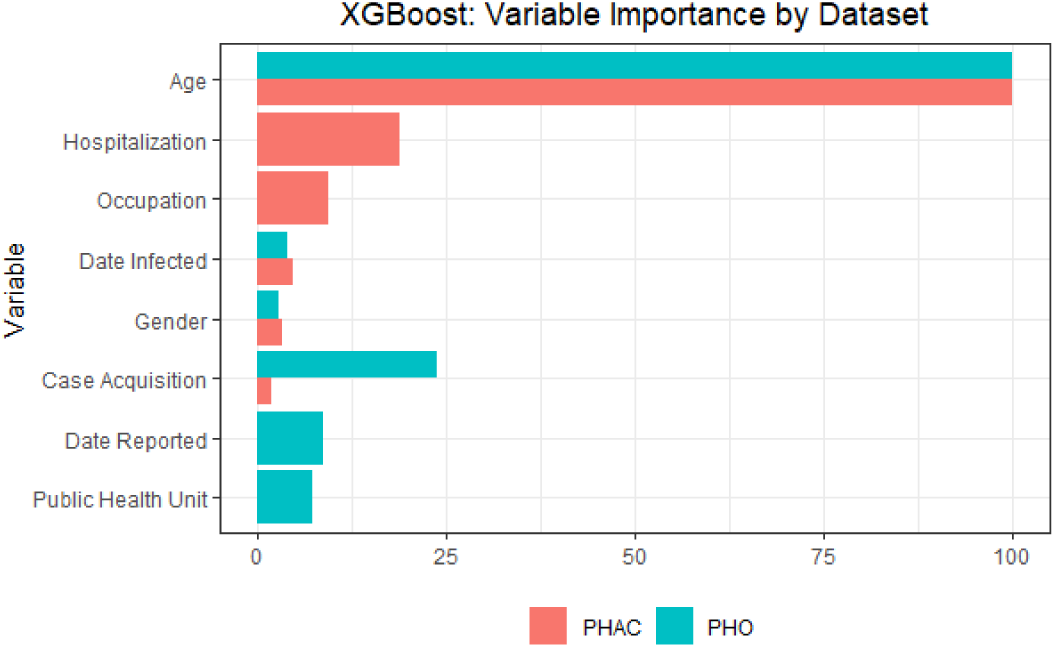
XGBoost Variable Importance graph by dataset.

Figure 3 depicts the variable importance for the XGBoost model for both datasets (PHO and PHAC). The variable importance is calculated as the gain associated with each variable for the final model, with the most important variable being scaled to 100. Age is demonstrated in Figure 3 to be the most important variable for the XGBoost model, for both datasets. The importance of the Age variable reflects the fact that long-term care facilities, where ages are typically high, experience much higher COVID-19 mortality risk (81% of deaths from COVID-19 in Canada occurred in Lon-Term Care homes. (CBC, 2020)).

The next two most important variables for the PHAC XGBoost model are Hospitalization and Occupation. Whether someone is hospitalized, or their occupation is a healthcare worker, has a substantial impact on predicting the mortality of a COVID-19 patient. This also indicates that the increased accuracy noticed for the models built using the PHAC dataset is likely in large part due to the addition of these two variables (hospitalization and occupation). The two variables located only in the PHO dataset, namely Date Reported and Public Health Unit, appear to be only relatively important for the model’s prediction. This indicates that by creating a publicly available dataset that contains all eight of these variables, improved accuracy of mortality prediction model would be attained.

### B. Differences in Early and Late Datasets

Comparing the mortality data between (i) the beginning of the COVID-19 outbreak and for the next four weeks during the ongoing pandemic, relative to (ii) the last five weeks data used a number of characteristics such as age, reveals how the demographics of the pandemic shifted over time. In ***Error! Reference source not found***. the cases and deaths reported in the first four weeks and last five weeks are contrasted. The number of deaths per confirmed case has dropped dramatically, from 0.047 fatalities per case in the winter to 0.0098 per case through the summer as evidenced in the data reports. Additionally, in the later period, all of the deaths were concentrated in the 80+ years and the ‘age not stated’ categories, while the earlier period recorded deaths in those as young as 50-59 years.

While the changes noted in Table 2 were influenced to some degree by the change in testing requirements which allowed the recording of a greater number of non-serious cases, the drop in mortalities also indicate there was considerable improvement (success) of treatment in hospitals. Specifically, as the pandemic progressed and many months have passed since COVID-19 was first reported in Canada, the global understanding of the disease, and improvements in treatment options occurred. Through research and medical practice, doctors in Canada now have more tools to fight serious cases of COVID-19 in hospitals. Steroids are now commonly used to significantly reduce deaths in patients with lung injuries as evidenced by studies from University of Oxford [32]. Additionally, anti-viral treatment Remedesivir has been approved in Canada to aid critically ill patients [32].

**TABLE 2.**
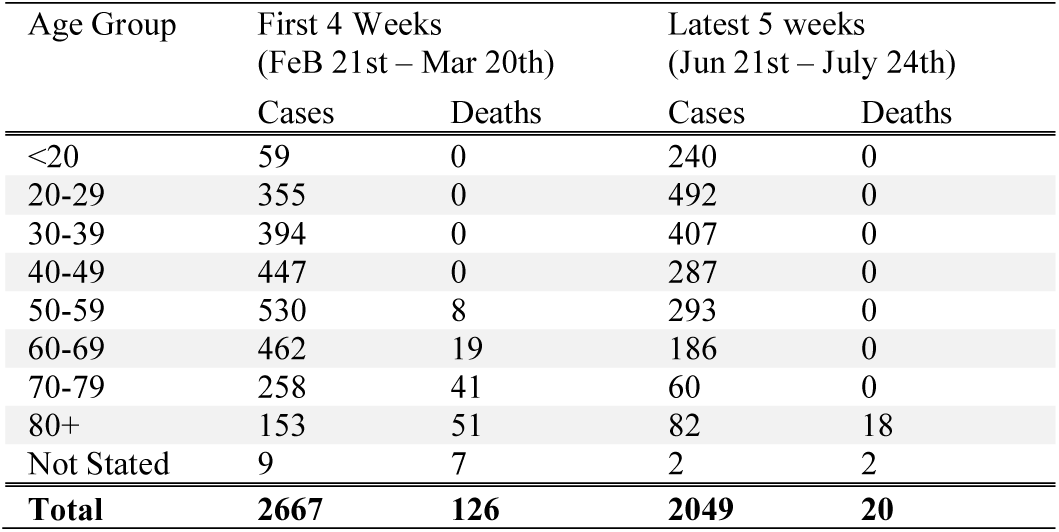
Comparison Between Earlier and Later Datasets.

Of interest in terms of exploring the power of the AI, training an XGBoost model on data from the two periods noted in Table 2, the results showed there was better accuracy, AUC, for the earlier time period of 0.914, and for the later time frame, the AUC fell to 0.838, but demonstrate good results for both. Most importantly, the AI results demonstrate the ability to detect the importance of different variables and detect trends (proven, as able to characterize the implications of the research and medical practices).

As indications of the adjustments identified over time, the results in Figure 4. XGBoost: Variable Importance by Time Period illustrate the changes in variable importance of the characteristics used by the XGBoost model are illustrated for the early and later datasets. The model based on the earlier data used a number of characteristics such as age, hospitalization status, date infected and gender to a much higher degree. The model based on later data relied almost entirely on age and somewhat on hospitalization, with minimal consideration for any of the other categories. As the results indicate in Table 2 showed, all the deaths were among elderly people (80+ years), indicating very clearly that age was identified as the very important issue for predicting mortality; the importance of hospitalization became dramatically less as a result of the learning curve from improved understanding, resulting in a dramatic decrease of importance of hospitalization resulting in mortality. These takeaways are, of course, now evident, from the reported results now available, but the key is that the AI model was able to identify the implications of the improved medical systems and understanding of the virus, to decrease the occasions of mortality.

**Figure 4.**
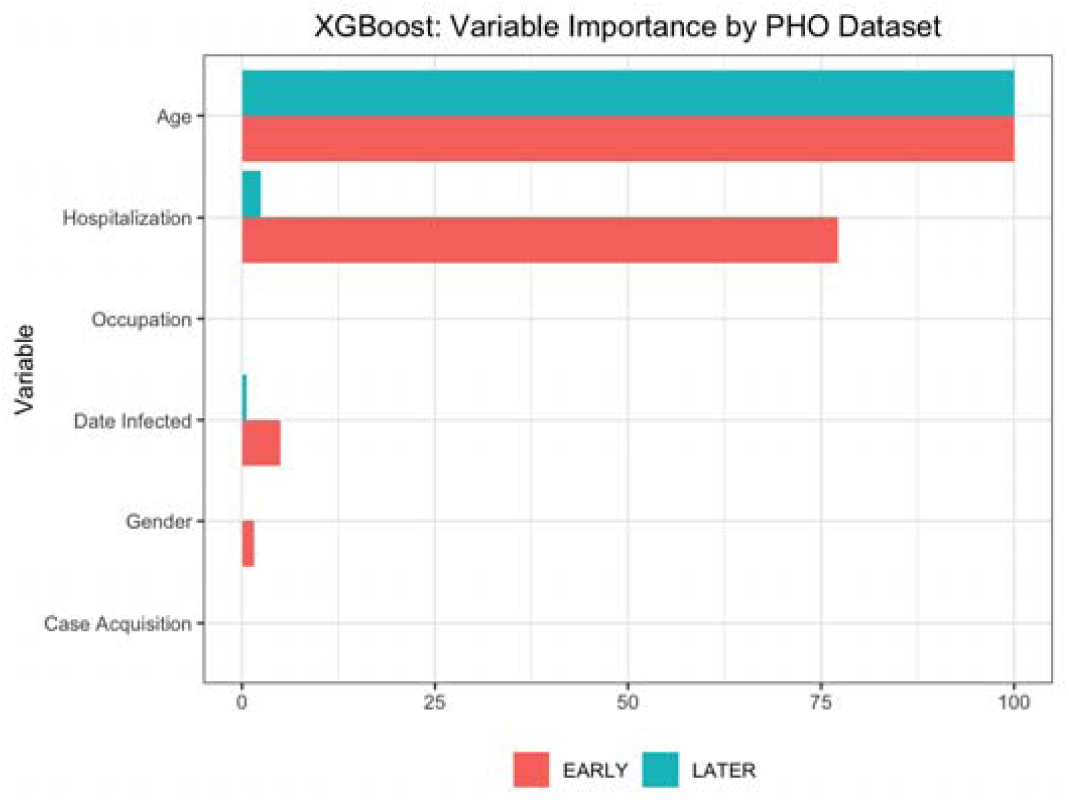
XGBoost: Variable Importance by Time Period.

**Figure 5.**
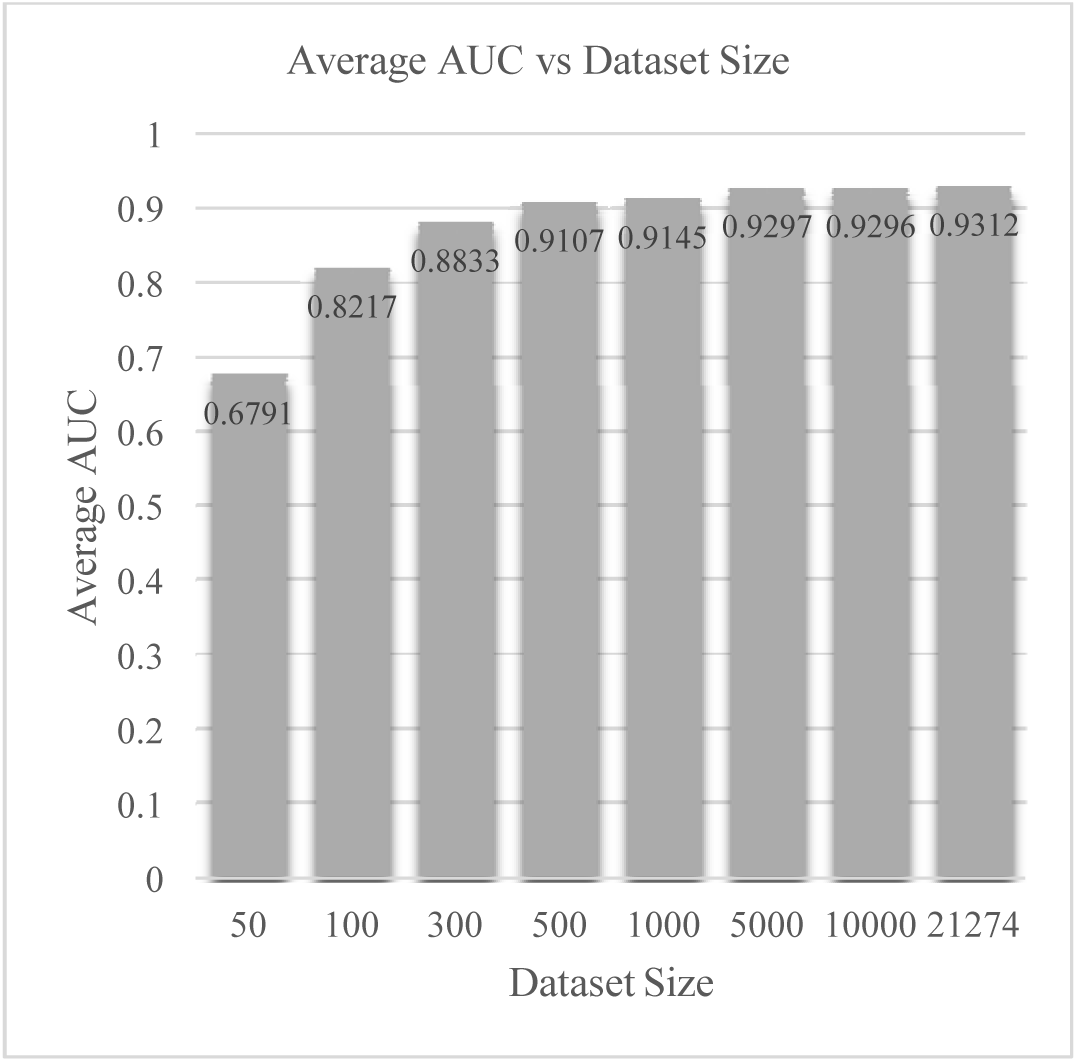
Average AUC versus Dataset Size.

### C. Number of Cases needed to build an accurate model

The accuracy of the models built in the previous sections were calibrated using a large training dataset containing over 27,000 patients. However, individual communities and hospitals may wish to develop their own COVID-19 mortality prediction model but may have significantly less cases available for AI model calibration. The following section identifies how the number of cases available for calibration may impact the models overall accuracy and thus provides guidance for these communities.

The impact of dataset size was analyzed using the most accurate model and dataset, XGboost – PHAC. Eight datasets with different sizes were created, using random sampling. A separate XGBoost model was trained on each dataset and the AUC was calculated using a hold-out testing dataset containing 30%of the overall Covid-19 case data. To ensure consistency, the entire process was repeated three times and the results averaged. Figure 4 below highlights the final averaged results for each dataset.

As expected, the AUC of the model increases with the size of dataset used to train the model. With only 50 cases to train the model the AUC is 0.6791 which indicates the model is not that effective in discerning the risk of mortality for COVID-19 patients. However, an XGBoost model trained using 300 cases has an AUC of 0.8833, indicating the model is fairly accurate and after the training dataset surpasses 1000 cases, the increase in AUC is minimal, only increasing by 0.0015 by including more than 10,000 additional cases into the training dataset.

### D. Discussion

From the three Machine Learning models and the one statistical model, the XGBoost model provides the highest level of precision and accuracy from both the PHO and PHAC datasets, as seen in ***Error! Reference source not found***.. This observation is critical to this study because it shows how an advanced machine learning model such as XGBoost, can provide more accurate COVID-19 outcomes in comparison to a statistical model, such as the logistics regression model.

In many respects, it is not surprising that the XGBoost performed well, as it has been a favourite in applied machine learning competitions since its release in 2014 due to its execution speed and model performance (e.g. [32] [33]).

Moreover, the results obtained using the national dataset (PHAC) are consistently higher than the results from the datasets release by the provincial government (PHO). The inconsistencies as to what information is made available between the two datasets are the cause of the different accuracy ranges, with key items being the inclusion of hospitalisation status and patient occupation in the PHAC database. However, the dataset available from PHO also produced significant results, with interesting attributes coming from the inclusion of location data pertaining to the Public Health Unit of the cases. The variable importance graph (Figure 3) suggests the PHAC results are bettered by taking into consideration hospital status as it displays the severity of the case; however, hospitalisation and ICU status may be useful for the mortality prediction of already severe COVID-19 cases, but this piece of data comes after the patient’s case has already reached a point of severity that requires medical intervention, and therefore cannot be used to predict whether or not a case will develop to the point of needing professional healthcare and possibly develop to the point of fatality.

AI has not yet been impactful against COVID-19 since the use of AI is hampered by availability of data related to issues of confidentiality to protect those and their families who have or may be impacted by this dreadful virus. There is much discussion about the mortality risk of COVID-19 patients in the province of Ontario as may be apparent through better data and AI modeling, to assess which patients to prioritize in receiving medical attention. Overcoming the constraints on data confidentiality will require a careful balance between data privacy and public health concerns, and more rigorous human-AI interaction.

## V. Conclusions

The implications of the Severe Acute Respiratory Syndrome COVID-19 virus (SARS-CoV-2) at world scale have been profound and the potential for initiation of a second wave of impacts from the virus are expected. The power of Artificial Intelligence (AI) as a technology was demonstrated as a means to improve prediction of the future impact on the medical care system. The needs for forecasting models are obvious, to provide the country’s leaders with information (e.g., how many people are likely going to be admitted to hospitals, and how many will require intensive care and/or ventilators, who is most at risk, etc.) to help them make informed decisions.

Substantial success was demonstrated as evident in terms of the accuracy of the XGBoost AI modeling, achieving an AUC of 0.9056 for PHO and 0.935 for the PHAC datasets. As well, the value of the AI modeling as gained from the importance of individual variables was demonstrated, providing important insights into the most important variable for the XGBoost model. Exploring the ability of the AI model to be sensitive to changing conditions, and in this case specifically, the improvements in the understanding how to treat the results, the AI model was able to identify the dramatic importance of the age of the mortality becomes dramatically important, with much lower impacts on the other variables that describe the individual caseloads. The results relying only upon the last five weeks of data showed that AI picks up the overwhelming importance of age is clearly evident, as an indication of mortality from COVID-19.

This is confirmed for the AI modelling wherein there was separate but very demonstrative evidence that showed long-term care facilities, where ages are typically high and comorbidities are frequent, are the most important factor differentiating between COVID-19 case outcomes (81% of deaths from COVID-19 in Canada occurred in Long-Term Care homes).

After age, the next two most important variables for the PHAC XGBoost model are Hospitalization and Occupation. Whether someone is hospitalized, or their occupation as a healthcare worker has a substantial impact on predicting the mortality of a COVID-19 patient. As well, the AI modeling showed that an XGBoost model trained using 300 cases has an AUC of 0.8833, indicating the model is fairly accurate, and that data after 1000 cases, improved accuracies from data are limited, indicating that development of an AI model from a much lesser basis than was the primary data utilized for this research, is still able to provide informative and useful results.

As the analyses described herein indicate, AI has the potential to be a tool in the fight against COVID-19 and similar pandemics. However, as Georgios Petropoulos concludes, “AI systems are still at a preliminary stage, and it will take time before the results of such AI measures are visible.” [34]

## Data Availability

The case data used in this manuscript are publicly available from Ontario Health and Statistics Canada websites

https://data.ontario.ca/dataset/f4112442-bdc8-45d2-be3c-12efae72fb27/resource/455fd63b-603d-4608-8216-7d8647f43350/download/conposcovidloc.csv

https://www150.statcan.gc.ca/t1/tbl1/en/tv.action?pid=1310078101

## Acknowledgment

Research funding by both the Natural Sciences Engineering Research Council (NSERC) under their special program for analyses of COVID-19 (ALLRP 549866-20) and by the University of Guelph Research Leadership Chair program are gratefully acknowledged.

Assistance in the ongoing research from Michael Chislett and Steven Yang are gratefully acknowledged as well as the many hardworking medical staff who have worked tirelessly on the front lines to respond to the ongoing crises and the many people who facilitated the data assembly by uploading the data to the various websites. All case data have been involved from publicly websites.

## References

[1] S. Ardabili, A. Mosavi, P. Ghamisi, F. Ferdinand, A. Varkonyi-oezy, U. Reuter, T. Rabezuk and P. Atkinson, “COVID-19 Outbreak Prediction with Machine Learning,” ResearchGate PrePrints, May 2020.

[2] W. Chen, H. R. Pourghasemi, A. Kornejady and N. Zhang, “Landslide spatial modeling: Introducing new ensembles of ANN, MaxEnt, and SVM machine learning techniques,” Geoderma, vol. 305, pp. 314–327, November 2017.

[3] M. Gumus and M. S. Kiran, “Crude oil price forecasting using XGBoost,” in 2017 International Conference on Computer Science and Engineering (UBMK), Antalya, 2017.

[4] F. Zhou, Q. Claire and R. D. King, “Predicting the Geographical Origin of Music,” in 2014 IEEE International Conference on Data Mining, Shenzhen, 2014.

[5] A. Shoukat, C. R. Wells, J. M. Langley, J. M. Singer, A. P. Galvani and S. M. Moghadas, “Projecting demand for critical care beds during COVID-19 outbreaks in Canada,” Canadian Medical Association Journal, vol. 192, no. 19, pp. E489-E496, 11 May 2020.

[6] J. Wiens and E. Shenoy, “Machine Learning for Healthcare: On the Verge of a Major Shift in Healthcare Epidemiology,” Clinical Infectious Diseases, pp. 149-153, 1 January 2018.

[7] J. Roth, M. Battegay, F. Juchler, J. Vogt and A. Widmer, “Introduction to Machine Learning in Digital Healthcare,” Infection Control & Hospital Epidemiology, vol. 39, no. 12, pp. 1457–1462, 2018.

[8] D. Jianga, M. Hao, F. Ding, J. Fu and M. Li, “Mapping the transmission risk of Zika virus using machine learning models,” Acta Tropica, pp. 391–399, 19 June 2018.

[9] S. Rose, “Mortality Risk Score Prediction in an Elderly Population Using Machine Learning,” American Journal of Epidemiology, vol. 177, no. 5, pp. 443–452, 01 March 2013.

[10] D. Chang, D. Chang and M. Pourhomayoun, “Risk Prediction of Critical Vital Signs for ICU Patients Using Recurrent Neural Network,” in 2019 International Conference on Computational Science and Computational Intelligence (CSCI), Las Vegas, 2019.

[11] M. Pourhomayoun, N. Alshurafa, B. Mortazavi, H. Ghasemzadeh, K. Sideris, B. Sadeghi, M. Ong, L. Evangelista, P. Romano, A. Auerbach, A. Kimchi and M. Sarrafzadeh, “Multiple model analytics for adverse event prediction in remote health monitoring systems,” in 2014 IEEE Healthcare Innovation Conference (HIC), Seattle, 2014.

[12] M. G. Chislett, P. Phillips, B. Snider, E. A. McBean, J. Yawney and S. A. Gadsden, “Assessing the Impact of Alternative Responses to COVID-19: Stopping the Spread in Newfoundland, Canada,” 2020.

[13] I. Holmdahl and C. Buckee, “Wrong but Useful — What Covid-19 Epidemiologic Models Can and Cannot Tell Us,” New England Journal of Medicine, vol. 383, no. 4, pp. 303–305, 23 July 2020.

[14] N. P. Dong, H. V. Long and A. Khastan, “Optimal control of a fractional order model for granular SEIR epidemic with uncertainty,” Communications in Nonlinear Science and Numerical Simulation, vol. 88, p. 105312, April 2020.

[15] X. Jiang, M. Coffee, A. Bari, J. Wang, X. Jiang, J. Huang, J. Shi, J. Dai, J. Cai, T. Zhang, Z. Wu, G. He and Y. Huang, “Towards an Artificial Intelligence Framework for Data-Driven Prediction of Coronavirus Clinical Severity,” Computers, Materials & Continua, vol. 62, no. 3, pp. 537–551, 30 March 2020.

[16] L. Yan, H.-T. Zhang, Y. Xiao, M. Wang, C. Sun, J. Liang, S. Li, M. Zhang, Y. Guo, Y. Xiao, X. Tang, H. Cao, X. Tan, N. Huang, B. Jiao, A. Luo, Z. Cao, H. Xu and Y. Yuan, “Prediction of criticality in patients with severe Covid-19 infection using three clinical features: a machine learning-based prognostic model with clinical data in Wuhan,” MedRxiv, 03 March 2020.

[17] B. Dickson, “AI is among our most effective tools in the fight against coronavirus,” TechTalks, 9 March 2020. [Online]. Available: https://bdtechtalks.com/2020/03/09/artificial-intelligence-covid-19-coronavirus/. [Accessed 10 August 2020].

[18] ttopstart, “Artificial Intelligence in Healthcare: Opportunities and Innovation Challenges,” [White paper], p. 3, 2 April 2020.

[19] Statistics Canada, “Health regions: boundaries and correspondence with census geography,” Government of Canada, 27 November 2015. [Online]. Available: https://www150.statcan.gc.ca/n1/pub/82-402-x/2011001/reg-eng.htm. [Accessed 14 August 2020].

[20] Public Health Ontario, “Enhanced Epidemiological Summary: COVID-19 in Long-Term Care Home Residents in Ontario,” Ontario Agency for Health Protection and Promotion, Toronto, 2020.

[21] Government of Ontario, “Confirmed positive cases of COVID-19 in Ontario,” 2020. [Online]. Available: https://data.ontario.ca/en/dataset/confirmed-positive-cases-of-covid-19-in-ontario/resource/455fd63b-603d-4608-8216-7d8647f43350.

[22] Government of Canada, “Detailed preliminary information on confirmed cases of COVID-19 (Revised),” 2020. [Online]. Available: https://www150.statcan.gc.ca/t1/tbl1/en/tv.action?pid=1310078101.

[23] Ontario, “Confirmed positive cases of COVID19 in Ontario,” 14 September 2020. [Online]. Available: https://www150.statcan.gc.ca/t1/tbl1/en/tv.action?pid=1310078101. [Accessed 14 September 2020].

[24] StatCan, “Detailed preliminary information on confirmed cases of COVID-19 (Revised), Public Health Agency of Canada,” August 2020. [Online]. Available: https://www150.statcan.gc.ca/t1/tbl1/en/tv.action?pid=1310078101. [Accessed 12 August 2020].

[25] W. Venables and B. Ripley, Modern Applied Statistics with S. Fourth Edition, New York: Springer. ISBN 0-387-95457-0, 2002.

[26] A. Liaw and M. Wiener, “Classification and Regression by RandomForest,” R News, vol. 2, no. ISSN 1609-3631, pp. 18–22, December 2002.

[27] T. Chen, T. He, M. Benesty, V. Khotilovi, Y. Tang, H. Cho, K. Chen, R. Mitchell, I. Cano, T. Zhou, M. Li, J. Xie, M. Lin, Y. Geng and Y. Li, “Version 1.0.0.2,” 2020. [Online]. Available: https://CRAN.R-project.org/package=xgboost.

[28] L. Torgo, “Data Mining with R, learning with case studies Chapman and Hall/CRC.,” 2010. [Online]. Available: http://www.dcc.fc.up.pt/~ltorgo/DataMiningWithR.

[29] R Core Team, “R: a language and environment for statistical computing,” Vienna, 2018.

[30] J. Huang and C. Ling, “Using AUC and Accuracy in Evaluating Learning Algorithms,” IEE Transactions on Knowledge and Data Engineering, vol. 17, no. 3, pp. 299–310, March 2005.

[31] C. Metz, “Basic principles of ROC analysis,” Seminars in Nuclear Medicine, vol. 8, no. 4, pp. 283–298, October 1978.

[32] J. Brownlee, “A Gentle Introduction to XGBoost for Applied Machine Learning,” MAchine Learning Mastery, 2016.

[33] E. A. M. Brett Snider, “Improving time to failure predictions for water distribution systems using extreme gradient boosting algorithm,” in Wdsa/ccwi joint conference proceedings, 2018.

[34] G. Petropoulos, “Artificial intelligence in the fight against COVID-19,” Bruegel, 23 March 2020.

[35] Q. Bi, Y. Wu, S. Mei, C. Ye, X. Zou, Z. Zhang, X. Liu, L. Wei, S. A. Truelove, T. Zhang, W. Gao, C. Cheng, X. Tang, X. Wu, Y. Wu, B. Sun, S. Huang, Z. Y. J. Sun and T. Ma, “Epidemiology and transmission of COVID-19 in 391 cases,” The Lancet Infectious Diseases, 27 April 2020.

[36] N. Ogden, A. Fazil, J. Arino, P. Berthiaume, D. Fisman, A. Greer, A. Ludwig, V. Ng and A. Tuite, “Modelling scenarios of the epidemic of COVID-19 in Canada,” CCDR, 04 June 2020.

[37] A. Tuite, D. Fisman and A. Greer, “Mathematical modelling of COVID-19 transmission and mitigation strategies in the population of Ontario, Canada,” CMAJ Research, 11 May 2020.

[38] T. Alamo, D. Reina and P. Millan, Data-Driven Methods to Monitor, Model, Forecast and Control Covid-19 Pandemic: Leveraging Data Science, Epidemiology and Control Theory, Cornell University COVID-19 e-prints, 2020.

[39] CSSE, “COVID-19 Dashboard,” 30 July 2020. [Online]. Available: https://gisanddata.maps.arcgis.com/apps/opsdashboard/index.html#/bda7594740fd40299423467b48e9ecf6.

[40] E. Dong, H. Du and L. Gardner, “An interactive web-based dashboard to track COVID-19 in real time,” The Lancet Infectious Diseases, vol. 20, no. 5, pp. 533–534, 1 May 2020.

[41] Centers for Disease Control and Prevention, “Nonpharmaceutical Interventions (NPIs),” U.S. Department of Health & Human Services, 27 April 2020. [Online]. Available: https://www.cdc.gov/nonpharmaceutical-interventions/index.html. [Accessed 5 August 2020].

[42] E. Alpaydin, Introduction to machine learning, 2nd ed., Cambridge, Massachusetts: MIT Press, 2010.

[43] S. Narkhede, “Understanding Confusion Matrix,” Towards Data Science, 2018.

[44] D. Fisman, A. Greer and A. Tuite, “Derivation and Validation of Clinical Prediction Rule for COVID-19 Mortality in Ontario, Canada,” MedRxiv, 2020.

[45] John Hopkins Coronavirus Resource Center, 2020. [Online]. Available: https://coronavirus.jhu.edu/map.html. [Accessed 14 September 2020].

[46] CBC News, “Canada’s proportion of COVID-19 deaths in long-term care double the average of other countries, study shows,” 25 June 2020. [Online]. Available: https://www.cbc.ca/news/health/coronavirus-canada-long-term-care-deaths-study-1.5626751. [Accessed 12 September 2020].

